# Titin’s role in diastolic heart function across species

**DOI:** 10.1101/2025.07.29.25332316

**Authors:** Antonio de Marvao, Rizwan Ahmed, Ronny Schweitzer, Kathryn A McGurk, Paolo Inglese, Pierre-Raphaël Schiratti, Soodeh Kalaie, Sean L Zheng, Saad Javed, Mit Shah, Chang Lu, Vladimir Losev, Deva Senevirathne, Lara Curran, Pantazis Theotokis, Rachel J Buchan, Wenjia Bai, Sebastian Schafer, Brian P Halliday, James S Ware, Stuart A Cook, Declan P O’Regan

**Affiliations:** MRC Laboratory of Medical Sciences, Imperial College London, Hammersmith Hospital Campus, London, UK; Department of Women and Children’s Health, King’s College London, London, UK; British Heart Foundation Centre of Research Excellence, School of Cardiovascular and Metabolic Medicine and Sciences, King’s College London, London, UK; National Heart and Lung Institute, Imperial College London, London, UK; Royal Brompton & Harefield Hospitals, Guy’s and St.Thomas’ NHS Foundation Trust, London, UK; Department of Computing, Imperial College London, UK; Department of Brain Sciences, Imperial College London, UK; National Heart Research Institute Singapore, National Heart Center Singapore, Singapore

## Abstract

**Background:** Cardiac diastolic function and blood pressure are physiologically tightly interconnected. While diastolic function is regulated by post-translational modifications of sarcomeric genes, including *titin* (*TTN*), isolating its genetic determinants from those influencing blood pressure is challenging due to variable *in vivo* loading conditions. We aimed to genetically map diastolic function to the rat genome to prioritise human gene orthologs for genotype-phenotype studies of cardiac function and clinical outcomes.

**Methods:** High-fidelity cardiac phenotyping using *ex vivo* Langendorff preparation in an F2 cross of inbred rat strains included evaluation of left ventricular contractility (LV) (dP / dt_max_) and relaxation (dP/dt_min_), while controlling loading conditions. In UK Biobank, whole exome sequencing was performed and cardiovascular image-derived phenotypes and outcomes were compared in adult rare variant carriers and genotype-negative participants.

**Results:** In rats, quantitative trait locus analysis identified a single locus on chromosome 3 that reached genome-wide significance for LV relaxation (dP/dt_min_), which was also associated with variation in LV contractility (dP/dt_max_) but not blood pressure. The *Ttn* gene, which encodes varied non-synonymous single-nucleotide polymorphisms of parental strains, is encoded at the peak of linkage. In humans, the prevalence of rare *titin* truncating variants (TTNtv) (allele frequency < 0.00004) in 454,756 UK Biobank participants was 0.37% (n = 1,672). Carrying a TTNtv was associated with an increased risk of death or major adverse cardiac events compared to genotype negative controls (hazard ratio: 1.94; 95% confidence interval [CI]: 1.78-2.12; *P* < 0.001), mainly due to heart failure endpoints (hazard ratio: 3.76; 95% CI: 3.30-4.29; *P* < 0.001). In 33,988 participants with both cardiac magnetic resonance imaging and whole exome sequencing, TTNtvs were associated with reduced diastolic function, even in participants with normal systolic function and no ventricular dilatation.

**Conclusions:** These studies demonstrate for the first time in rats, that naturally occurring variations in *Ttn* are associated with diastolic function, independently of blood pressure, and in humans TTNtv cause impaired cardiac relaxation and adverse outcomes before overt cardiomyopathy develops.

## Introduction

Our understanding of the genetic mechanisms underlying diastolic function primarily comes from loss-of-function, knock-out, and knock-in studies in murine models. These studies demonstrate that gene manipulation and consequent disruption of key sarcomeric proteins, such as myosin-binding protein C (Mybpc3)^1^, titin (Ttn)^2^, troponin T (Tnnt2) and troponin I (Tnni3),^3^ impairs myocardial relaxation and passive stiffness. However, there is limited direct genetic evidence from humans or other animals,^4^ and isolating the naturally occurring genetic variation that determines diastolic function remains challenging, as loading conditions are not easily controlled for *in vivo*. Blood pressure (BP) is intrinsically linked to diastolic function, as increased afterload and arterial stiffness impair ventricular relaxation and elevate filling pressures. This makes it difficult to discern whether loci identified in genome-wide association studies in humans primarily influence myocardial properties or act through haemodynamic pathways.^5^ Notably, while in rats BP regulation is mostly linked to quantitative trait locus (QTL) on chromosomes 1, 2, and 10, with chromosome 7 associated with salt-sensitive hypertension and chromosome 13 linked to the renin gene, in humans, BP QTLs are distributed across all chromosomes.^6^ *Ex vivo* perfused rat heart models offer a unique advantage in this context, enabling precise control of preload and afterload to assess intrinsic myocardial properties and provide a platform for exploring the genetic regulation of diastolic function.

Here we aimed to understand the role of natural genetic variation in modulating diastolic function using phenotyping of a rodent model. We investigated a genome-sequenced F2 intercross from inbred rat strains that exhibit phenotypic uniformity in physiological experiments and enable association of these phenotypes with refined regions of the genome while adjusting for covariates such as BP. To date, there has been no reported animal data on genetic determination of left ventricular (LV) myocardial relaxation (dP/dt_min_) or myocardial contractility (dP/dt_max_). We then sought to further assess these associations on diastolic function in participants from UK Biobank (UKB) as well as lifetime risk of adverse events. Using high-precision, deep-learning phenotyping of cardiac magnetic resonance imaging (CMR), we characterized phenotypic manifestations of systolic and diastolic function. Together, these experiments across species allow the identification of candidate genes for BP-independent effects on diastolic function and evaluating their phenotypic expression in populations.

## Methods & Materials

### Rodent methods

Genome-sequenced inbred rat strains Brown Norway (BN) and spontaneously hypertensive rat (SHR) were used to perform high-fidelity *ex vivo* cardiovascular phenotyping. We proceeded an F2 intercross from BN and SHR. Rats were bred by a monogamous mating system. BN females were crossed with SHR males to produce BN X SHR (BXH) F1 animals and a reciprocal cross was performed to obtain SHR x BN (HXB) animals. F1 BXH animals were intercrossed to produce F2 BXH animals and F1 HXB animals were intercrossed to produce F2 HXB animals. Cardiovascular phenotyping was undertaken in 184 F2 rats. Animals were maintained in the Central Biomedical Services facility, Imperial College London and housed at a maximum of five per cage. All procedures were performed in accordance with the UK Animals (Scientific Procedures) Act of 1986.

Blood pressure was measured using cannulation of the carotid artery in each animal before cardiac excision for *ex vivo* phenotyping. Animals were anaesthetized using inhaled 4% Isoflurane. Haemodynamic data was captured continuously by LabchartPro software (ADI instruments, Dunedin, New Zealand) and analyzed offline.

Following BP measurement, depth of anaesthesia was confirmed before cardiac excision and immediate arrest in ice-cold Kreb’s buffer to cause cardiac standstill before transfer to *ex vivo* perfusion apparatus. Hearts were gently mounted onto the aortic cannula via the aorta. Perfusion was established within a couple of minutes of cardiac excision. The left atrium (LA) was then removed, and a fluid-filled latex balloon was placed in the LV cavity. LV contractility (LV dP/dt_max_) and LV relaxation (LV dP/dt_min_) were derived from LV pressure. A co-axial bipolar electrode was placed on the right atrium and pacing was commenced at 360 beats per minute. The whole preparation was kept inside a jacketed container which was maintained at 37 ^°^ C. The isolated heart preparation was studied at baseline for 15 minutes. Myocardial infarction was subsequently induced by ligation of the proximal left anterior descending (LAD) artery for 35 minutes and this was followed by reperfusion for an hour. Cardiac mass was measured at the end of the procedure.

DNA was extracted from the tissue or blood using robotic extraction on the Maxwell16 system (Promega, Madison, WI). High throughput genotyping was performed using Illumina’s (San Diego, CA) GolgenGate assay mapping 768 single nucleotide polymorphisms (SNP) in 184 F2 individuals. QTL mapping was undertaken using the R/qtl package.^7^ The logarithm of odds (LOD) score is a log-likelihood of the probability of a QTL at a locus given the phenotype values to there being no QTL. Permutation tests demonstrated that in our dataset there was less than a 5% probability of encountering a LOD threshold greater than 3.9 across the entire genome (Supplementary Figure 1). This was taken as LOD threshold for a significant QTL.

### UK Biobank study

The UKB study recruited over 500,000 participants aged 40 to 69 years old from across the United Kingdom between 2006 and 2010 (National Research Ethics Service, 11/NW/0382).^8^ This study was conducted under terms of access approval number 40616 and 47602. Written informed consent was provided. A sub-study of UKB invited participants for CMR assessment of cardiac chamber volumes and function using a standard protocol (1.5T, Siemens Aera, Siemens Medical Systems, Erlangen, Germany).^9^ We selected 33,988 participants who underwent whole exome sequencing (WES) and did not harbor any pathogenic / likely pathogenic (P/LP) variants in cardiac genes associated with dilated cardiomyopathy (DCM) other than *TTN*. A summary of the analysis is shown in Figure 1.

**Figure 1.**
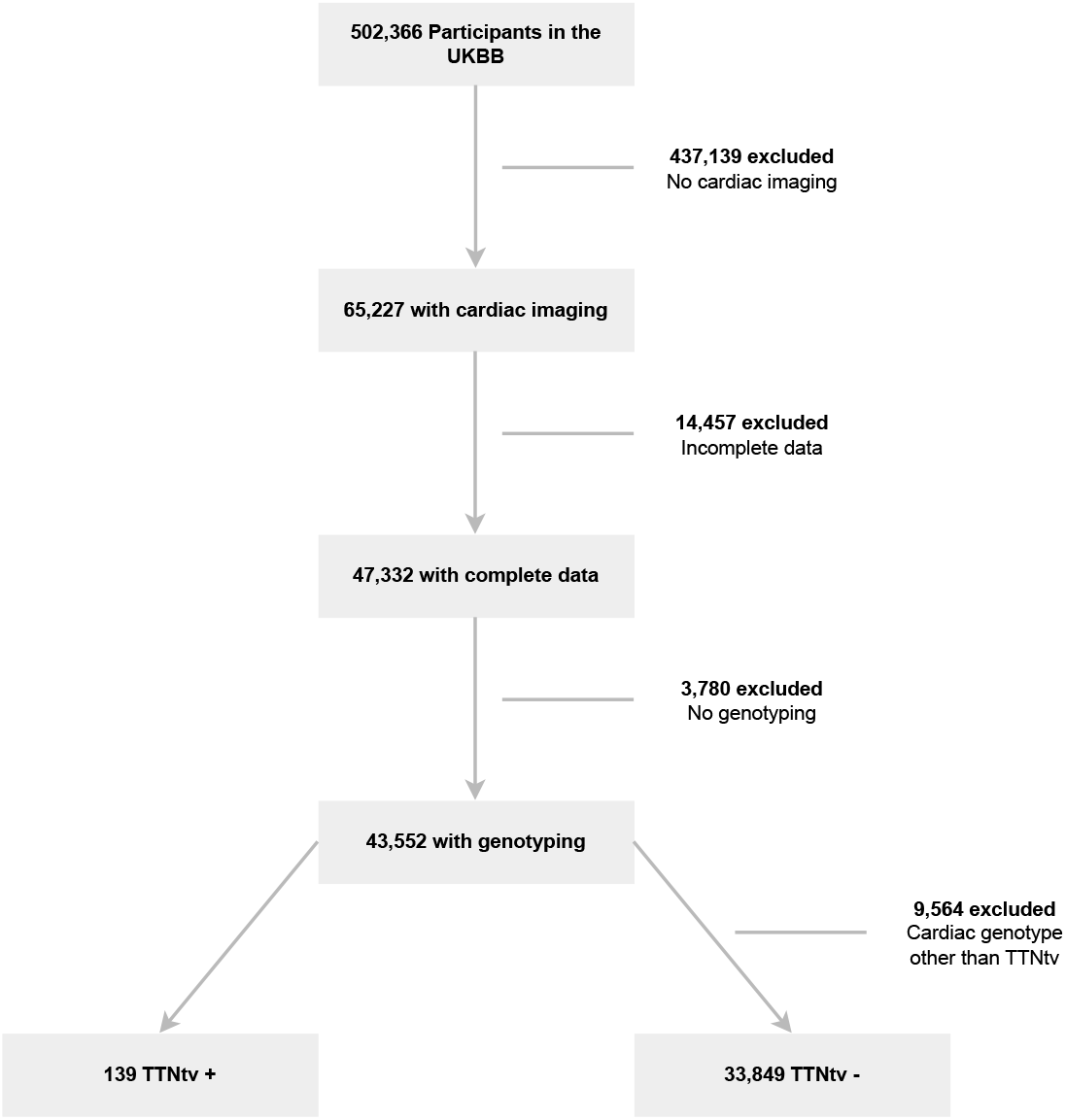
Study flowchart. Details of participants included in the study and reasons for exclusion. UKBB, UK Biobank; TTNtv, *Titin*-truncating variants.

### Human cardiac phenotyping using machine learning

Segmentation of cine images in UKB was performed using a deep learning neural network algorithm developed and optimised in-house. The performance of image annotation using this algorithm is equivalent to a consensus of expert human readers and achieves sub-pixel accuracy for cardiac segmentation.^10^ The label maps were super-resolved and registered to a cardiac atlas enabling consistent quantitative three-dimensional phenotypic analysis within and between patient groups.^11^

Myocardial wall thickness was measured along radial line segments connecting the endocardial and epicardial surfaces perpendicular to the myocardial centreline and excluding trabeculae. Chamber volumes and mass were calculated from the segmentations according to standard post-processing guidelines.^12^ The association between genotype and three-dimensional phenotype was assessed by fitting univariable regression models at each vertex of the cardiac mesh, controlling for false discovery, with corrected beta coefficients plotted on the epicardial surface.^13,14^

Myocardial strain analysis was performed using non-rigid free-form deformation image registration between successive frames (Figure 2).^15^ To reduce the accumulation of registration errors motion tracking was performed in both forward and backward directions from the end-diastolic (ED) frame and an average displacement field calculated.^5^ Radial (*E*_rr_) strains were calculated on the short axis cines by the change in length of respective line segments. Motion tracking was also performed on the long-axis four-chamber cines to derive longitudinal (*E*_ll_) strain. Peak strain for each segment and global peak strain were then calculated. Strain was measured from slices acquired at basal, mid-ventricular, and apical levels. Strain rate was estimated as the first derivative of strain and peak early diastolic strain rate in radial (PDSR_rr_) and longitudinal (PDSR_ll_) directions was detected using an algorithm to identify local maxima.

**Figure 2.**
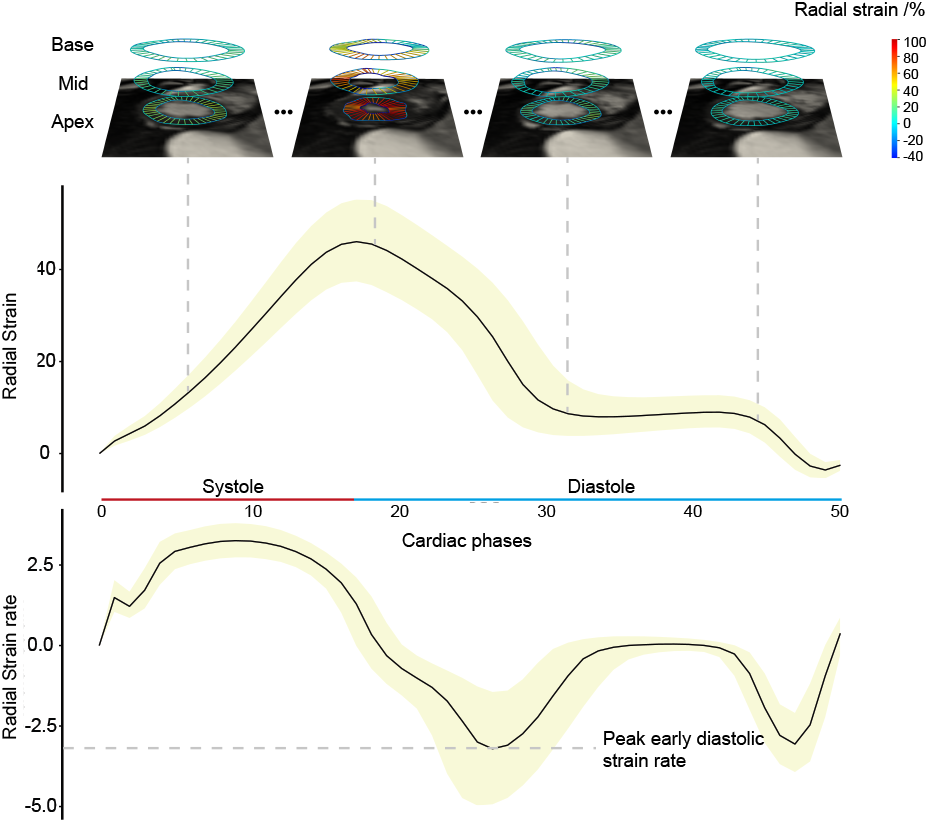
Machine learning analysis of cardiac imaging. Cardiac magnetic resonance imaging was segmented using convolutional neural networks followed by motion analysis using the deformation between cine frames. Peak early diastolic strain rate was used as a marker of diastolic function and was calculated from the first derivative of left ventricular strain. Based on Thanaj *et al*.^5^ Population median values with inter-quartile ranges shown.

### Variant categorization pipeline

UKB participants underwent WES as previously described.^16^ Individuals were classified as genotype negative if they did not carry a rare protein-altering genetic variation (minor allele frequency (MAF) < 0.001 in the UKB and the Genome Aggregation Database (gnomAD v2.1))^17^ in 19 DCM-associated genes with definitive, strong, or moderate evidence for disease. Variants were extracted from the WES data of 454,756 UKB participants. Matched Annotation from NCBI and EMBL-EBI (MANE), protein-altering variants (PAVs) that had a MAF < 0.00004% in gnomAD and UKB were identified. Only TTNtvs in ‘percent spliced in’ (PSI) ratio > 90% exons predicted to influence gene product level (e.g. frameshift, essential splice, stop gained) were kept. Compound heterozygous carriers of common TTNtvs (same two *TTN* variants identified in > 10 individuals) were removed from the analysis. The variant list only included variants that would be called P/LP if identified in a patient with DCM; using Cardioclassifier and ClinVar, and the variants were manually curated if they had any evidence of pathogenicity.

### Outcome measures

The effect of genotype status on clinical outcomes was assessed by using lifetime risk. The UKB reports the date of first occurrence of a diagnosis, identified from self-reporting, primary care, hospital in-patient, and death register records. This permitted the identification of events preceding recruitment to the UKB. The primary clinical outcome was a composite of all-cause mortality or major adverse cardiovascular events (MACE) defined as a diagnostic code for heart failure (including cardiomyopathy), arrhythmia, stroke, or cardiac arrest events. Secondary clinical outcomes were the individual components of the primary clinical outcome. A full list of endpoint definitions and data fields used from the UKB database is presented in the (Supplementary Table 1).

### Statistical Analysis

Statistical analysis was performed with R version 4.2.2 (R Foundation for Statistical Computing), unless otherwise stated. Variables are expressed as percentages if categorical, mean ± standard deviation (SD) if continuous and normal, and median (interquartile range) if continuous and non-normal. Baseline anthropometric data were compared by a Wilcoxon test. Imaging parameters were compared by using analysis of covariance, adjusted for relevant clinical covariates.

Clinical outcomes were analyzed in participants with WES stratified according to genotype categories. Cox proportional hazards were calculated for lifetime risk of clinical events. For the primary outcome, competing risk analysis was performed by using the cause-specific survival method. Secondary analysis of incident clinical events from recruitment excluded individuals with events preceding enrollment and was performed by using Cox proportional hazards adjusted for age at recruitment. Time-to-event was censored at first event for each outcome, death, or last recorded follow-up. The relationship between genotype or CMR phenotypes and outcomes was assessed with multivariable Cox proportional hazards models. Sex was included as a covariate in all models. Hazard proportionality assumption was tested by using Schoenfeld residuals. Sex was found to be in violation of these assumptions, and therefore a sex-stratified analysis was conducted with interaction coefficients. Outcomes are reported as hazard ratios (HRs) with 95% confidence intervals (CIs) and presented graphically as cumulative hazards and Cox proportional hazards curves.

Mass univariate analysis was performed on the local phenotypes of the three-dimensional representation of the left ventricle at ED and ES of 19,185 participants. Local wall thickness and geometric differences were adjusted for age, sex, body surface area (BSA) and systolic BP using TTNtv status as a predictor.

In all cases statistical significance was considered at *P* ≤ 0.05.

## Results

### Rodent models

Single QTL analysis for cardiovascular traits at baseline revealed a common genetic locus for LV dP/dt_min_ and LV dP/dt_max_ on rat chromosome 3 (Figure 3). The QTL was statistically significant with a LOD score of 9.3 for LV dP/dt_min_ and 7 for LV dP/dt_max_ with genome-wide significance *P* < 0.001. The QTL has a large effect size and accounts for 17.2% (369 ± 75 mmHg/s) of the variation in LV dP/dt_max_ and 21.8% (310 ± 54 mmHg/s) of the variation seen in LV dP/dt_min_. QTL significance thresholds and LOD scores remain unaffected after taking into account systolic and diastolic BP on LV dP/dt_min_ (Supplementary Figures 2 and 3). Inheritance of the BN allele, encoding multiple non-synonymous single-nucleotide polymorphisms (nsSNPs), was associated with lower LV dP/dt_max_ and higher LV dP/dt_min_ values (Supplementary Figure 4). The coding region polymorphisms in the *Ttn* sequence between BN and SHR are shown in Supplementary Table 2.

**Figure 3.**
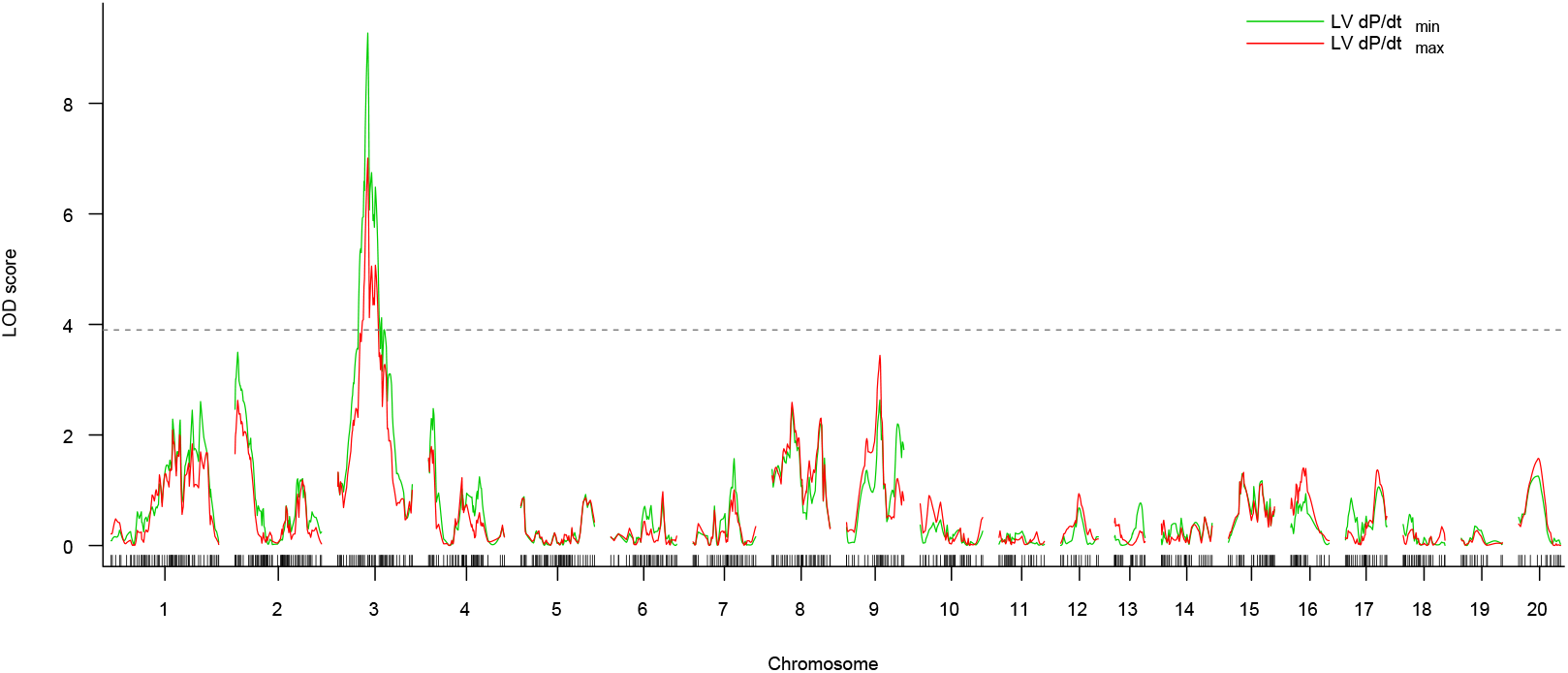
Quantitative trait loci in rats. Genome-wide LOD plot for myocardial relaxation (LV dP/dt_min_) and myocardial contractility (LV dP/dt_max_) showing identical linkage peak on chromosome 3. Vertical bars on the x-axis represent genotyped markers on each chromosome.

### Prevalence of variants in humans

A summary of participant characteristics is shown in Table 1. The prevalence of rare TTNtvs (allele frequency < 0.00004) in 454,756 participants was 0.37% (n = 672/453,084). In the subgroup with imaging 139 had TTNtv, while 9,564 participants with imaging harbored other pathogenic/likely pathogenic variants in cardiac genes and were excluded from analysis, resulting in 33,849 genotype negative participants.

**Table 1.**
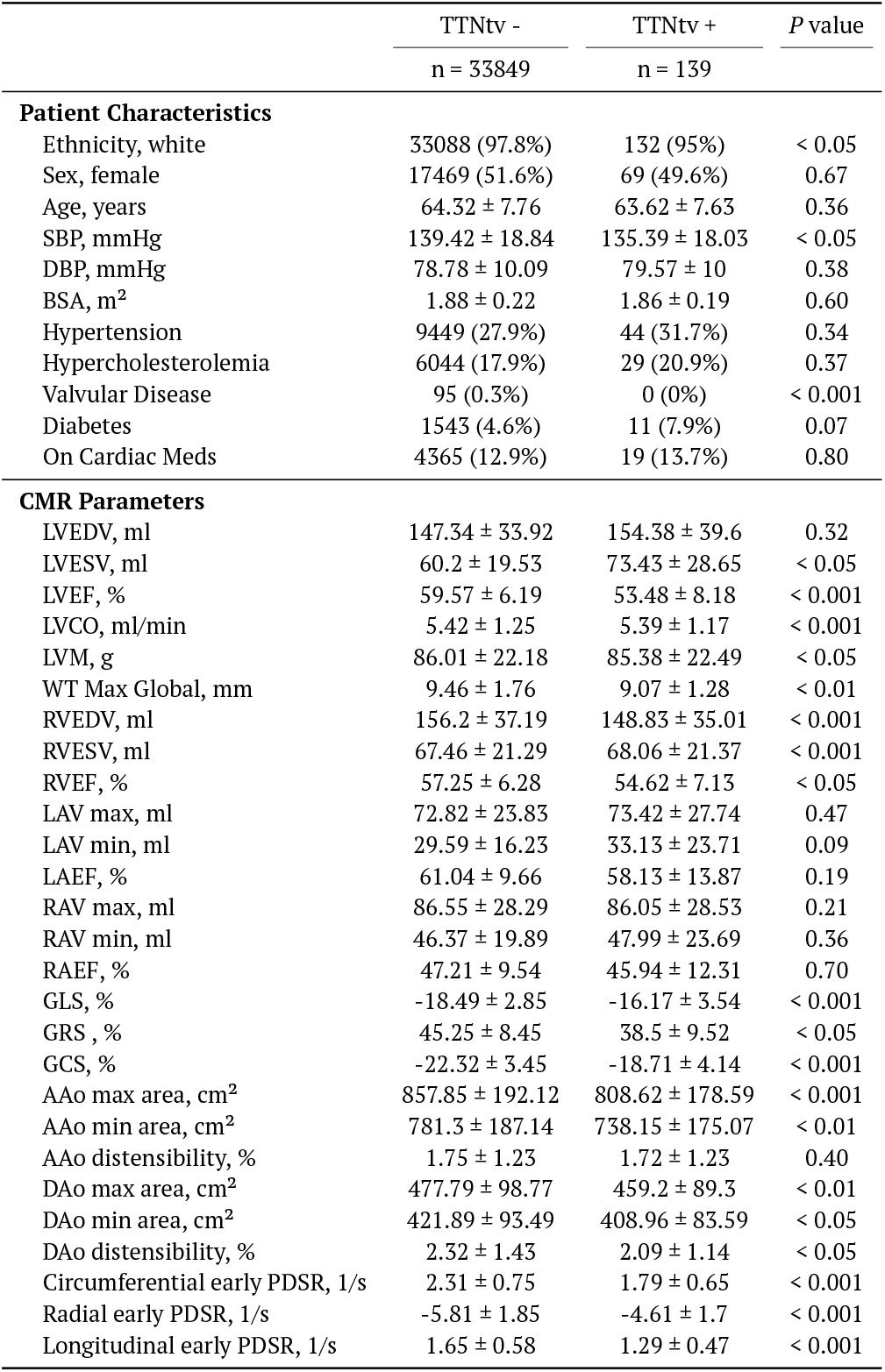
Patient characteristics and CMR-derived measurements by genotype. Categorical data - number of patients (relative frequency); Continuous data - mean ± standard deviation. BSA, body surface area; CMR, cardiac magnetic resonance imaging; DBP, diastolic blood pressure; EDV, end-diastolic volume; EF, ejection fraction; ESV, end-systolic volume; LA, left atrial; LV, left ventricular; LVM, left ventricular mass; RA, right atrial; RV, right ventricular; SBP, systolic blood pressure; WT, wall thickness; LVEDV, left ventricular end-diastolic volume; LVESV, left ventricular end-systolic volume; LVCO, left ventricular cardiac output; LVEF, left ventricular ejection fraction; RVEDV, right ventricular end-diastolic volume; RVESV, right ventricular end-systolic volume; RVEF, right ventricular ejection fraction; LAV, left atrial volume; LAEF, left atrial ejection fraction; RAV, right atrial volume; RAEF, right atrial ejection fraction; AAo, ascending aorta; DAo, descending aorta; PDSR, peak diastolic strain rate; GLS, global longitudinal strain; GRS, global radial strain; GCS, global circumferential strain.

|  | TTNtv -<br>n = 33849 | TTNtv +<br>n = 139 | P value |
| --- | --- | --- | --- |
| **Patient Characteristics** |  |  |  |
| Ethnicity, white | 33088 (97.8%) | 132 (95%) | < 0.05 |
| Sex, female | 17469 (51.6%) | 69 (49.6%) | 0.67 |
| Age, years | 64.32 ± 7.76 | 63.62 ± 7.63 | 0.36 |
| SBP, mmHg | 139.42 ± 18.84 | 135.39 ± 18.03 | < 0.05 |
| DBP, mmHg | 78.78 ± 10.09 | 79.57 ± 10 | 0.38 |
| BSA, m<sup>2</sup> | 1.88 ± 0.22 | 1.86 ± 0.19 | 0.60 |
| Hypertension | 9449 (27.9%) | 44 (31.7%) | 0.34 |
| Hypercholesterolemia | 6044 (17.9%) | 29 (20.9%) | 0.37 |
| Valvular Disease | 95 (0.3%) | 0 (0%) | < 0.001 |
| Diabetes | 1543 (4.6%) | 11 (7.9%) | 0.07 |
| On Cardiac Meds | 4365 (12.9%) | 19 (13.7%) | 0.80 |
| **CMR Parameters** |  |  |  |
| LVEDV, ml | 147.34 ± 33.92 | 154.38 ± 39.6 | 0.32 |
| LVESV, ml | 60.2 ± 19.53 | 73.43 ± 28.65 | < 0.05 |
| LVEF, % | 59.57 ± 6.19 | 53.48 ± 8.18 | < 0.001 |
| LVCO, ml/min | 5.42 ± 1.25 | 5.39 ± 1.17 | < 0.001 |
| LVM, g | 86.01 ± 22.18 | 85.38 ± 22.49 | < 0.05 |
| WT Max Global, mm | 9.46 ± 1.76 | 9.07 ± 1.28 | < 0.01 |
| RVEDV, ml | 156.2 ± 37.19 | 148.83 ± 35.01 | < 0.001 |
| RVESV, ml | 67.46 ± 21.29 | 68.06 ± 21.37 | < 0.001 |
| RVEF, % | 57.25 ± 6.28 | 54.62 ± 7.13 | < 0.05 |
| LAV max, ml | 72.82 ± 23.83 | 73.42 ± 27.74 | 0.47 |
| LAV min, ml | 29.59 ± 16.23 | 33.13 ± 23.71 | 0.09 |
| LAEF, % | 61.04 ± 9.66 | 58.13 ± 13.87 | 0.19 |
| RAV max, ml | 86.55 ± 28.29 | 86.05 ± 28.53 | 0.21 |
| RAV min, ml | 46.37 ± 19.89 | 47.99 ± 23.69 | 0.36 |
| RAEF, % | 47.21 ± 9.54 | 45.94 ± 12.31 | 0.70 |
| GLS, % | -18.49 ± 2.85 | -16.17 ± 3.54 | < 0.001 |
| GRS, % | 45.25 ± 8.45 | 38.5 ± 9.52 | < 0.05 |
| GCS, % | -22.32 ± 3.45 | -18.71 ± 4.14 | < 0.001 |
| AAo max area, cm<sup>2</sup> | 857.85 ± 192.12 | 808.62 ± 178.59 | < 0.001 |
| AAo min area, cm<sup>2</sup> | 781.3 ± 187.14 | 738.15 ± 175.07 | < 0.01 |
| AAo distensibility, % | 1.75 ± 1.23 | 1.72 ± 1.23 | 0.40 |
| DAo max area, cm<sup>2</sup> | 477.79 ± 98.77 | 459.2 ± 89.3 | < 0.01 |
| DAo min area, cm<sup>2</sup> | 421.89 ± 93.49 | 408.96 ± 83.59 | < 0.05 |
| DAo distensibility, % | 2.32 ± 1.43 | 2.09 ± 1.14 | < 0.05 |
| Circumferential early PDSR, 1/s | 2.31 ± 0.75 | 1.79 ± 0.65 | < 0.001 |
| Radial early PDSR, 1/s | -5.81 ± 1.85 | -4.61 ± 1.7 | < 0.001 |
| Longitudinal early PDSR, 1/s | 1.65 ± 0.58 | 1.29 ± 0.47 | < 0.001 |

Among the TTNtv+ group (n = 139), the mean age was 64 ± 8 years, 95% were of British ancestry, and 49.6% female. In comparison, the TTNtv-group (n = 33,849) had a mean age of 64 ± 8 years, 97.8% were British ancestry, and 51.6% female. There were no significant differences in age (*P* = 0.36) or sex (*P* = 0.67), though there was a significant difference in ancestry (*P* < 0.05).

### Human diastolic function

Participants with TTNtv had a higher end-systolic volume (LVESV) (73.43 ± 28.65 ml) compared to genotype-negative controls (60.2 ± 19.53 ml) (*P* < 0.05). However, there was no significant difference in ED volume (LVEDV; 154.38 ± 39.6 ml vs 147.34 ± 33.92 ml; *P* = 0.32). The ejection fraction (LVEF) was significantly lower in the TTNtv group (53.48 ± 8.18%) compared to the negative genotype group (59.57 ± 6.19%; *P* < 0.001).

When compared to genotype-negative participants, those carrying TTNtv exhibited lower global longitudinal strain (−16.17 ± 3.54% vs −18.49 ± 2.85%; *P* < 0.001), lower global circumferential strain (−18.71 ± 4.14% vs −22.32 ± 3.45%; *P* < 0.001), and lower global radial strain (38.5 ± 9.52% vs 45.25 ± 8.45%; *P* < 0.05) (Figure 4). When comparing early PDSR, a measure of diastolic function, the TTNtv group had lower rates in all directional measures compared to genotype-negative participants: circumferential PDSR (1.79 ± 0.65 1/s vs. 2.31 ± 0.75 1/s, *P* < 0.001), radial early PDSR (−4.61 ± 1.7 1/s vs. −5.81 ± 1.85 1/s, *P* < 0.001), longitudinal early PDSR (1.29 ± 0.47 1/s vs. 1.65 ± 0.58 1/s, *P* < 0.001). Findings were similar when restricting the analysis to participants with normal left ventricular ejection fraction (LVEF >50%) as shown in Supplementary Table 3.

**Figure 4.**
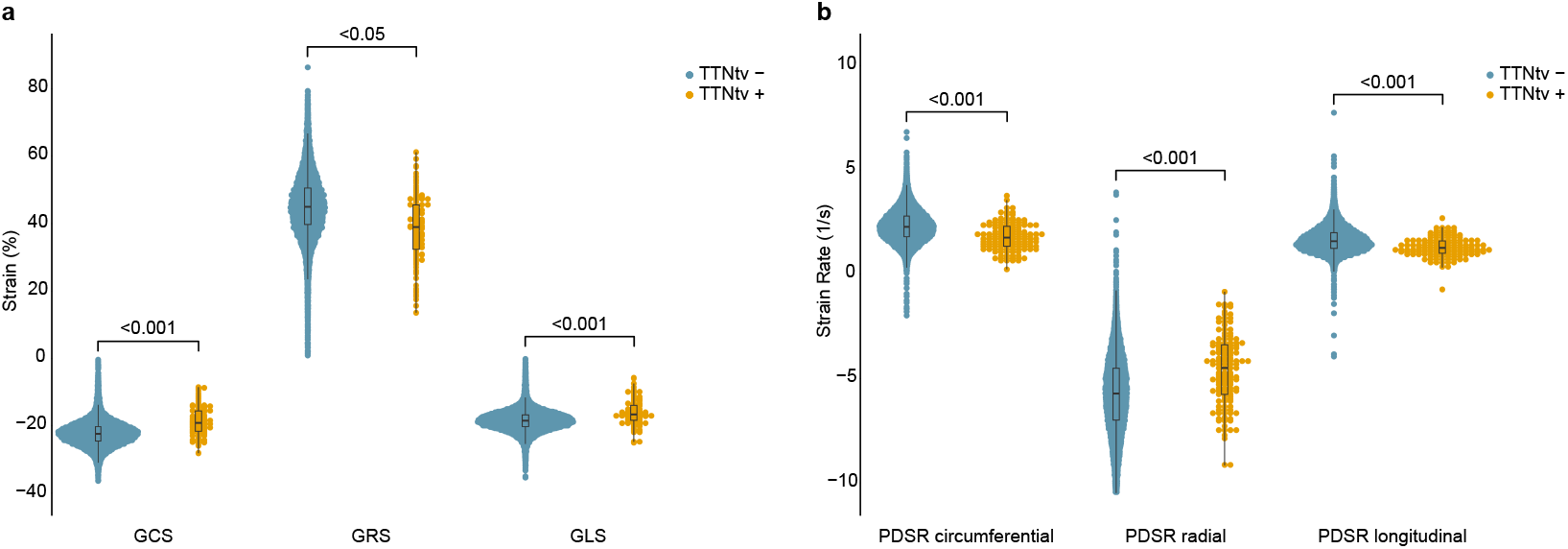
Strain data in humans. Comparison of **a.**systolic strain and **b**. diastolic strain rates by genotype. Pairwise *P* value comparisons shown. GCS, global circumferential strain; GRS, global radial strain; GLS, global longitudinal strain; PDSR, peak early diastolic strain rate, TTNtv, *Titin*-truncating variants.

### Remodelling in humans

The three-dimensional local phenotypes of the participants were analysed with mass univariate linear regression to evaluate associations with TTNtv carrier status. At ED the septal and lateral walls showed the greatest outward remodelling in TTNtv carriers, while at ES the entire LV cavity was enlarged (Figure 5a). These results align with the two-dimensional findings showing higher LVESV. There were no differences in regional wall thickness for TTNtv carriers (Figure 5b). We found that TTNtv carriers showed non-uniform changes in LV motion compared to controls with the strongest effects localised to the basal septal region (Supplementary Figure 5).

**Figure 5.**
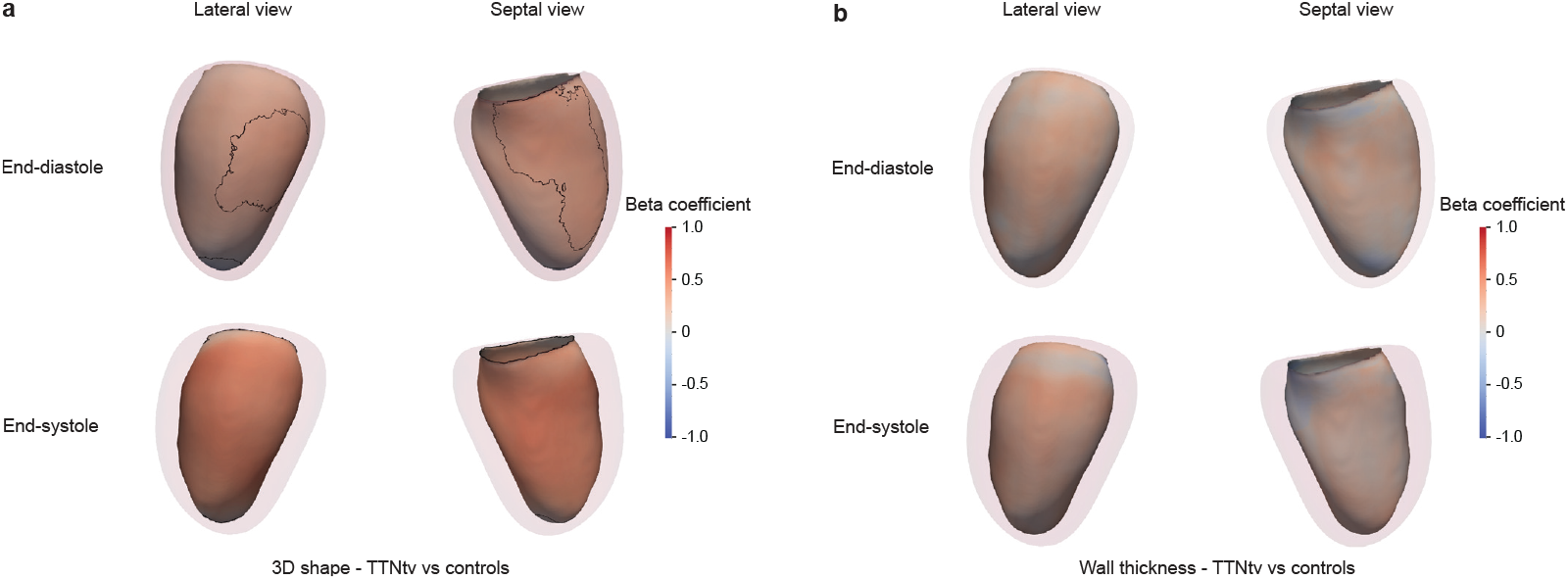
Mass univariate analysis of remodelling phenotypes using three-dimensional models of the left ventricle. **a.**Mass univariate linear regression was performed on local distance measurements showing a positive beta coefficient where TTNtv carrier status is associated with greater regional expansion. Sample size was 19,195 participants. The endocardium is represented from septal and lateral projections along with an outline of the epicardium. Black outlines represent significant areas of differences between genotype status *P* ≤ 0.05. For end-systole, the whole left ventricle is significant for geometric effects. **b**. The same mass univariate linear regression performed on local wall thickness. Here, no area is significant for TTNtv carrier status on wall thickness.

### Clinical outcomes

Clinical outcomes for 502,247 participants were analyzed (Figure 6). Median age at imaging was 65 years (interquartile range: 58-70 years), and participants were followed up for a median of 14.7 years (interquartile range: 14.0-15.5 years) with a total of 128,524 individual clinical events reported. Among genotype negative participants (n = 500,576), and those harbouring TTNtv (n = 1,671) individuals, there were 88,921 (17.8%) and 510 (30.5%) primary clinical outcome events (all-cause mortality or MACE), and by age 70 years there were 41,030 (cumulative incidence: 10.46%) and 276 events (cumulative incidence: 20.45%), respectively.

**Figure 6.**
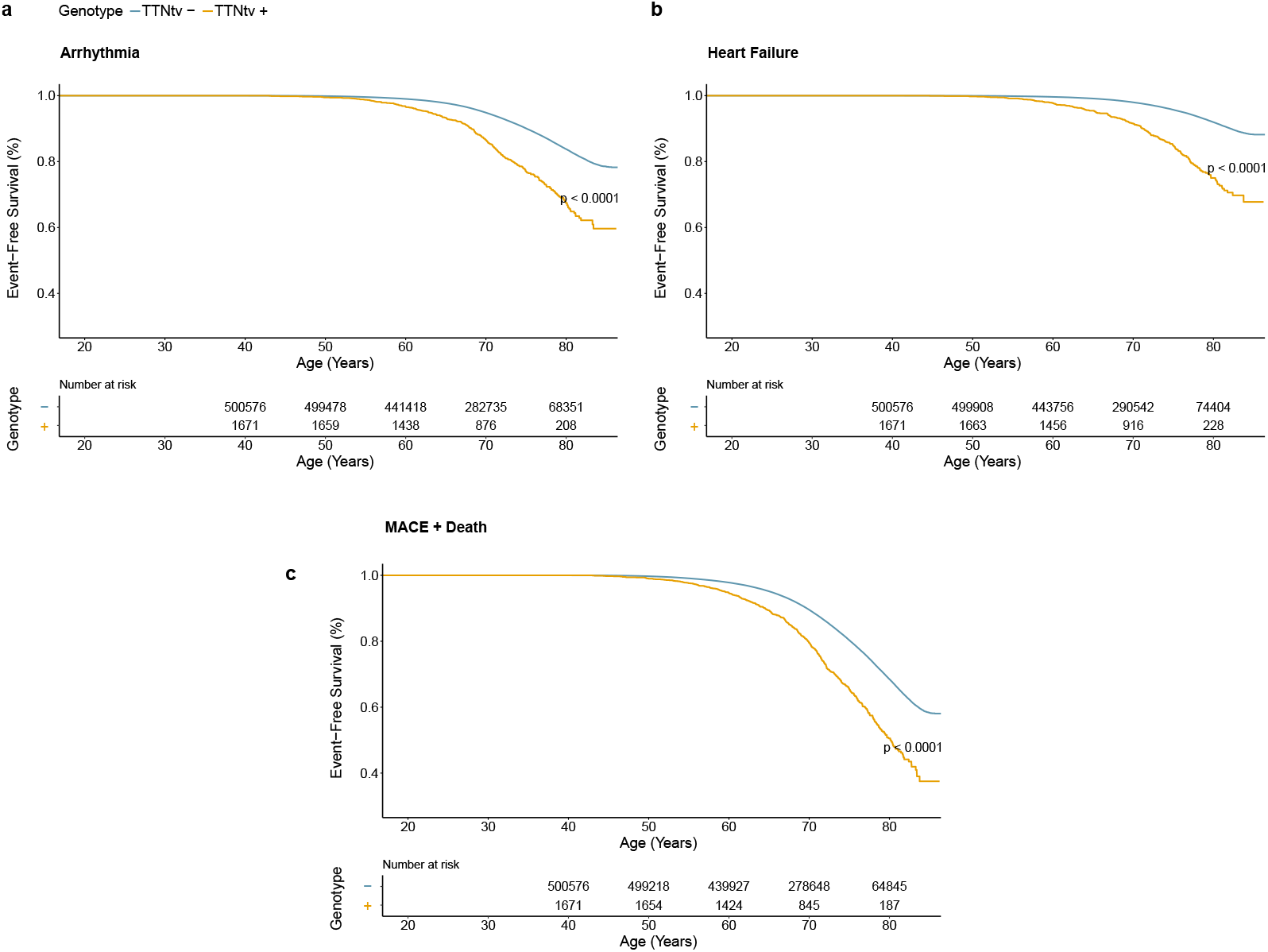
Kaplan-Meier plots showing the event-free survival for clinical events. **a.**Arrhythmia events, **b**. Heart failure events, **c**.MACE and death events. TTNtv, *Titin*-truncating variants; MACE, major adverse cardiovascular events.

Examining lifetime risks, TTNtv variants were associated with an increased risk of death or MACE (HR: 1.94; 95% CI: 1.78-2.12; *P* < 0.001), heart failure (HR: 3.76; 95% CI: 3.30-4.29; *P* < 0.001), and arrhythmia (HR: 2.43; 95% CI: 2.18-2.72; *P* < 0.001). Sensitivity analyses of lifetime risk of death or MACE, excluding participants with any cardiomyopathy and excluding cardiomyopathy events as an outcome, yielded concordant results to the primary analysis comparing genotype negative participants with those harboring TTNtv (excluding participants with cardiomyopathy HR: 1.89 [95% CI: 1.73-2.01]; *P* < 0.001; Figure 7; excluding cardiomyopathy diagnosis from the HF composite outcome HR: 1.89 [95% CI: 1.73-2.07]; *P* < 0.001; *P* < 0.001), confirming that previous findings were not solely driven by individuals with known disease, nor only by a diagnostic label of cardiomyopathy as an endpoint.

**Figure 7.**
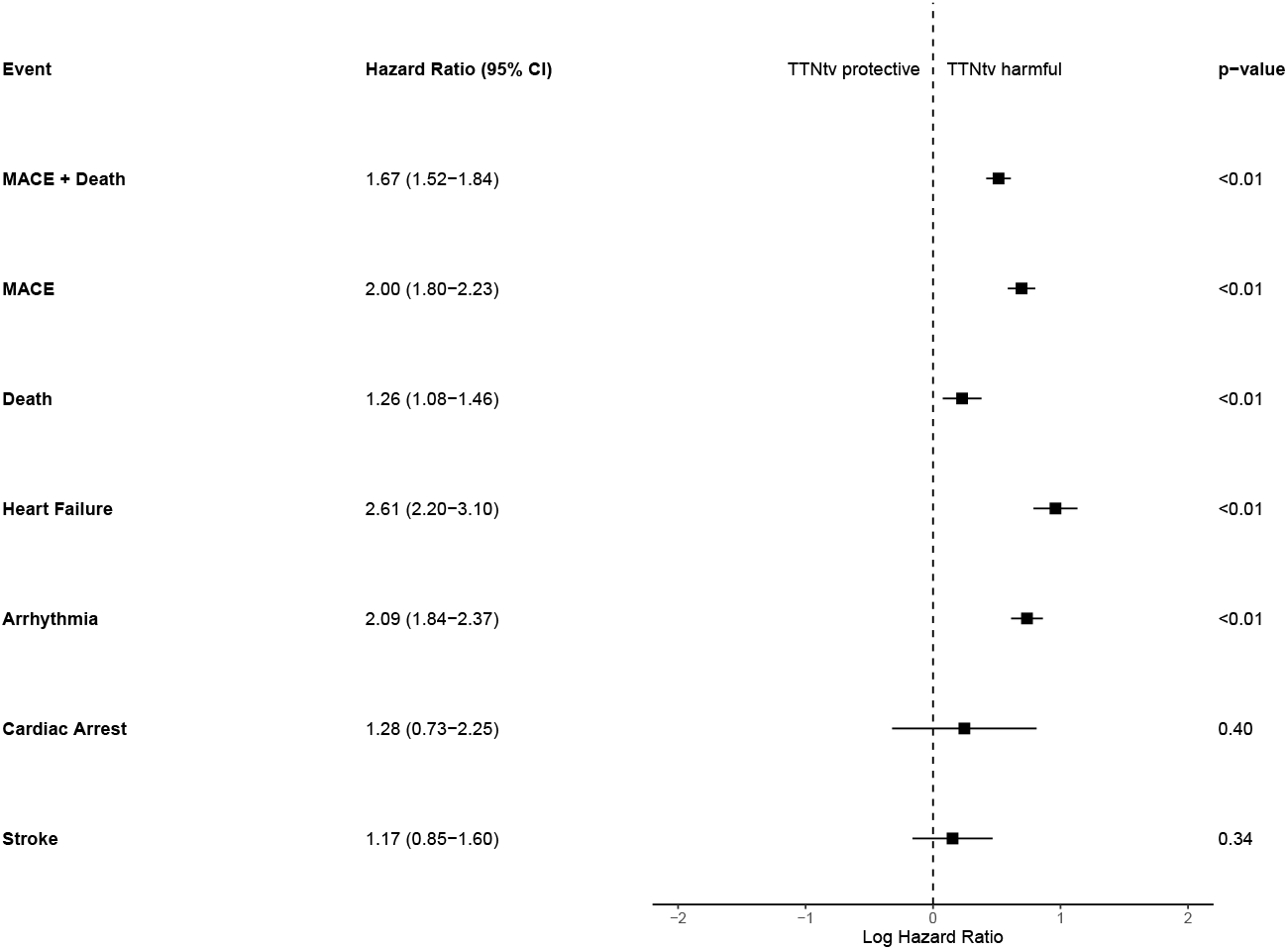
Forest plot showing the hazard ratios for clinical endpoints according to TTNtv status. Participants with a clinical diagnosis of cardiomyopathy excluded. *Titin*-truncating variants; MACE, major adverse cardiovascular events.

## Discussion

Through phenotyping of an animal model we determined that naturally occurring variants in *Ttn* modulate diastolic function independently of BP, consistent with a sarcomeric ‘spring’ effect during contraction and relaxation.^18^ We then showed in an adult human population that TTNtv are associated with diastolic function - with or without a history of cardiomyopathy. Genetic manipulation of *Ttn* is known to impair active and passive myocyte properties and here we demonstrate for the first time the effects on diastolic function of naturally occurring variation across species.

Our initial search for plausible biological candidates in the relatively small linkage region for LV dP/dt_min_ and LV dP/dt_max_ on rat chromosome 3 identified two potential candidate genes, *Ttn* and *Mybp3*, both highly expressed in cardiac tissue. However, *Mybp3* does not have any coding region polymorphisms between the two rat strains studied, leaving *Ttn* as the only potential candidate. *Ttn* also lies in proximity to the QTL peak whereas *Mybp3* lies at the fringe of the QTL interval. Titin extends from the I band region of the sarcomere to the M band. There are two descriptive sections comprising the I-band, which is elastic and extensible, and the A-band which is inextensible. I-band titin has three isoforms N2B, N2BA (composed of N2B and an additional N2A region) and a fetal isoform. N2B and N2BA isoforms are both expressed in adult hearts with N2BA having a longer extensible region and being more compliant than the N2B isoform; the ratio of the two isoforms may determine passive stiffness in adult mammalian heart.^19,20^ Phosphorylation at serine residues in the N2B segment additionally reduces titin-dependent passive tension.^19^ A comparison of the rat *Ttn* sequence variation between BN and SHR revealed the presence of nine nsSNPs in the SHR genome compared with the reference BN genome. Five of these nsSNPs are present in exon 44 which codes for part of the N2B sequence, and two nsSNPs result in the replacement of the serine residues, one at position 3905, resulting in the replacement of serine with phenylalanine, and the other at position 4138 replacing serine with leucine. These substitutions are predicted to affect protein function based on alteration in amino acid properties.^21^

These data from animal studies demonstrate myocardial relaxation as a hertitable quantitative trait segregating indepen-dently of BP and myocardial mass to a QTL that harbors *Ttn* close to peak linkage. Furthermore, genetic variation between the two strains studied postulate the role of the N2B isoforms or phosphorylation at these sites to modulate myocardial stiffness. The giant protein titin interconnects the thin and thick filaments within cardiac sarcomeres, serving as a structural scaffold, signaling hub, and source of passive tension and elasticity in muscle cells.^18^ Experimental interventions in previous mouse models, including domain deletion studies, have demonstrated that altering *Ttn* impairs diastolic function.^22^ However, evidence for a similar effect of naturally occurring variation is limited in both rats and humans. Variants in this gene play an important role in the pathogenesis of DCM, a condition characterized by decreased myocardial contractility and ventricular dilation.^23–26^ *Titin* truncating variants may mediate the development of DCM through a dual mechanism of proteinopathy and haploinsufficiency although the pathogenic mechanisms are incompletely understood.^27–29^

We then assessed the functional and prognostic consequences of TTNtv in a middle-aged adult population where 1 in 270 individuals harbored variants in exons predicted to influence gene product levels. We found that participants showed an increase in ES volumes and a lower ejection fraction, while ED volumes were unaffected as previously reported.^30^ Using motion analysis we further found that systolic strain was reduced in both short and long axis directions. The PDSR, a measure of the rate of contractile relaxation during diastole, was lower even when excluding those with an ejection fraction > 50%. Three-dimensional phenotyping showed that regional remodelling was present in ED but that the changes in geometry were stronger and more widespread at ES. The prevalence of primary outcomes by the age of 70 was almost twice that of genotype negative controls. In terms of lifetime risk, TTNtv were predominantly associated with a higher risk of heart failure and arrhythmia even when excluding those with known cardiomyopathy or reduced systolic function. These findings show that although TTNtv are associated with an attenuated cardiomyopathic phenotype individuals who harbor these variants have an increased risk of adverse cardiovascular events and exhibit diastolic dysfunction.

In conclusion, we show that naturally occurring genetic variation in *TTN* independently contributes to diastolic function across species with loss-of-function variants associated with impaired cardiac relaxation, remodelling and adverse outcomes.

## Data Availability

Data from UK Biobank are available for approved research. Other data are available upon reasonable request to the authors.

## Author contributions

A.de M. and R.A. performed data curation and analysis, and drafted the manuscript; R.A. performed analysis of the animal data; K.A.McG., L.C., R.J.B. and P.T. performed the genetic analysis; P.I., M.S., R.S., L.C., V.L. and P-R.S. performed analysis and visualisation of the data; A. de M. and W.B. performed the image analysis; S.L.Z. performed the survival analysis; B.P.H. and S.S. contributed to the drafting to the manuscript; S.A.C., J.S.W. and D.P.O’R. conceived the project, provided supervision and funding. All authors contributed to the article and approved the submitted version.

## Funding

The study was supported by Medical Research Council (MC_UP_1605/13); National Institute for Health Research (NIHR) Imperial College Biomedical Research Centre; the British Heart Foundation (RG/F/24/110138, RE/24/130023, CH/F/24/90015, FS/IPBSRF/22/27059, FS/ICRF/21/26019, NH/F/23/70013), Engineering and Physical Sciences Research Council (EP/W01842X/1, EP/Z531297/1), Rosetrees Trust and the Fetal Medicine Foundation (495237). D.P.O’R., S.A.C and J.S.W. are supported by the British Heart Foundation’s Big Beat Challenge award to CureHeart (BBC/F/21/220106).

## Data availability

Population data used in this study are available from UK Biobank. Other data are available upon reasonable request to the authors.

## Disclosures

J.S.W. has received research support from BMS, has acted as a paid advisor to Health Lumen, Tenaya Therapeutics, and Solid Biosciences, and is a founder with equity in Saturnus Bio. D.P.O’R has consulted for Bayer AG and BMS, and receives grant support from Bayer AG and Calico Labs. S.A.C. is co-founder and shareholder of Enleofen Bio Pte Ltd and VVB Bio Pte Ltd. B.P.H. has consulted for AZ and Eli Lily. The remaining authors have nothing to disclose.

## Notes

### Author Declarations

The UK National Research Ethics Service gave ethical approval for the UK Biobank study (11/NW/0382).

