## Supplementary Material for "Titin’s role in diastolic heart function across species"

### Supplementary Tables 1 - 3

| Endpoint | Definition | UKB data-field ID(s) | ICD-10 Code(s) |
|---|---|---|---|
| *Patient Characteristics* | | | |
| Ethnicity | Self-reported Ethnicity | 21000 | N/A |
| Sex | Biological Sex at Birth | 31 | N/A |
| Age | Age at Imaging Visit | 12697 | N/A |
| SBP | Systolic Blood Pressure | 4080, 93 | N/A |
| DBP | Diastolic Blood Pressure | 4079, 94 | N/A |
| BSA | Body Surface Area | Derived from 21002 (weight), 50 (height) | N/A |
| Hypertension | Diagnosed Hypertension | 131286, 6150, 41202, 41204 | I10-I15 |
| Hypercholesterolemia | Elevated cholesterol diagnosis | 6153, 6177, 41202, 41204 | E78.0, E78.5 |
| Valvular Disease | Any Valvular Pathology | 41202, 41204 | I34-I39 |
| Diabetes | Type 1 or Type 2 diabetes | 2443, 41202, 41204 | E10, E11 |
| On Cardiac Meds | Current cardiac medication use | 6153, 6177, 20003 | N/A |
| *Clinical Outcomes* | | | |
| All-cause Mortality | Death from any Cause | 40000 (date), 40001 / 40002 (cause) | Any |
| Heart failure | First Heart Failure / Cardiomyopathy Record | 131298, 41202, 41204, 42018 | I50, I42 |
| Arrhythmia (incl. AF) | First Arrhythmia Event | 131424, 41202, 41204, 42018 | I47-I49, I48 |
| Stroke | First Ischaemic or Haemorrhagic Stroke | 131040, 41202, 41204, 42018 | I60-I63 |
| Cardiac arrest | First Sudden Cardiac arrest / Ventricular stand-still | 41202, 41204, 40000 | I46 |
| MACE | Composite of HF, Arrhythmia, Stroke or Arrest | 131040, 131298, 131424, 41202, 41204, 42018 | I42, I46, I47-I49, I48, I50, I60-I63 |
| MACE+Death | MACE or All-cause Death | 131040, 131298, 131424, 40000, 41202, 41204, 42018 | I42, I46, I47-I49, I48, I50, I60-I63, or any cause of death |
| *CMR-Derived Variables* | | | |
| CMR variables | All Variables derived from Cardiac MRI images | 20207 | N/A |

**Supplementary Table 1. UK Biobank study variable definitions, data-field IDs, and ICD-10 codes for cardiovascular outcomes analysis.** AF, atrial fibrillation; BSA, body surface area; CMR, cardiac magnetic resonance; DBP, diastolic blood pressure; HF, heart failure; ICD-10, International Classification of Diseases 10th Revision; MACE, major adverse cardiovascular events (composite of heart failure, arrhythmia, stroke, or cardiac arrest); N/A, not applicable; SBP, systolic blood pressure; UKB, UK Biobank.

| SNP position | Exon | Reference AA | Alternate AA | AA position |
|---|---|---|---|---|
| chr3:59661015 | 3 | A | V | 121 |
| chr3:59654243 | 6 | T | A | 394 |
| chr3:59600175 | 44 | L | S | 3864 |
| chr3:59600095 | 44 | D | N | 3891 |
| chr3:59600052 | 44 | S | F | 3905 |
| chr3:59599359 | 44 | V | A | 4136 |
| chr3:59599353 | 44 | S | L | 4138 |
| chr3:59468374 | 117 | A | T | 9405 |
| chr3:59468146 | 118 | L | P | 9450 |
| chr3:59464765 | 123 | V | I | 9939 |

**Supplementary Table 2. Consequences of nsSNPs in the Titin sequence between BN and SHR.** Exon 44 codes for part of the N2B sequence of Titin and previous studies have shown that phosphorylation occuring at Serine residues in the N2B results in diastolic function improvement[31]. Two of the five mutations in exon 44 result in replacement of the Serine (S) residues by F (Phenylalanine) and L (Leucine). Others include V (Valine), A (Alanine), T (Threonine), D (Aspartate), I (Isoleucine), L (Leucine), N (Asparagine) and P (Proline).

| | TTNtv - | TTNtv + | p-value |
|---|---|---|---|
| | n = 31866 | n = 98 | |
| **Patient Characteristics** | | | |
| Ethnicity, white | 31145 (97.7%) | 91 (92.9%) | < 0.01 |
| Sex, female | 17018 (53.4%) | 59 (60.2%) | 0.19 |
| Age, years | 64.19 ± 7.73 | 62.94 ± 7.84 | 0.16 |
| SBP, mmHg | 78.62 ± 10.01 | 78.49 ± 9.83 | 0.85 |
| DBP, mmHg | 139.36 ± 18.84 | 135.06 ± 17.9 | < 0.05 |
| BSA, $m^2$ | 1.87 ± 0.22 | 1.82 ± 0.18 | < 0.05 |
| Hypertension | 8717 (27.4%) | 26 (26.5%) | 0.91 |
| Hypercholesterolemia | 5570 (17.5%) | 19 (19.4%) | 0.60 |
| Valvular Disease | 78 (0.2%) | 0 (0%) | < 0.001 |
| Diabetes | 1369 (4.3%) | 8 (8.2%) | 0.07 |
| On Cardiac Meds | 3843 (12.1%) | 10 (10.2%) | 0.76 |
| **CMR Parameters** | | | |
| LVEDV, ml | 145.95 ± 32.59 | 142.55 ± 31.74 | 0.19 |
| LVESV, ml | 58.22 ± 16.71 | 60.69 ± 16.83 | 0.15 |
| LVEF, % | 60.4 ± 5.13 | 57.68 ± 4.82 | < 0.001 |
| LVCO, ml/min | 5.45 ± 1.25 | 5.44 ± 1.19 | < 0.001 |
| LVM, g | 85.07 ± 21.61 | 78.66 ± 18.01 | < 0.001 |
| WT Max Global, mm | 9.41 ± 1.73 | 8.92 ± 1.26 | 0.16 |
| RVEDV, ml | 155.46 ± 36.94 | 143.84 ± 32.85 | < 0.001 |
| RVESV, ml | 66.3 ± 20.52 | 61.7 ± 18.26 | < 0.001 |
| RVEF, % | 57.77 ± 5.7 | 57.41 ± 5.27 | < 0.05 |
| LAV max, ml | 72.14 ± 22.82 | 66.61 ± 17.81 | 0.16 |
| LAV min, ml | 28.77 ± 14.43 | 25.41 ± 10.1 | 0.08 |
| LAEF, % | 61.56 ± 9 | 62.81 ± 8.03 | 0.08 |
| RAV max, ml | 85.78 ± 27.33 | 80.04 ± 23.86 | 0.07 |
| RAV min, ml | 45.56 ± 18.55 | 40.67 ± 15.24 | < 0.01 |
| RAEF, % | 47.55 ± 9.29 | 49.78 ± 10.49 | < 0.01 |
| GLS, % | -18.7 ± 2.65 | -17.43 ± 2.74 | < 0.001 |
| GRS, % | 45.98 ± 7.88 | 42.5 ± 7.3 | 0.30 |
| GCS, % | -22.69 ± 3.05 | -20.68 ± 2.82 | < 0.001 |
| AAo max area, $cm^2$ | 853.43 ± 190.12 | 785.9 ± 158.08 | < 0.01 |
| AAo min area, $cm^2$ | 776.93 ± 185.25 | 716.01 ± 157.53 | < 0.05 |
| AAo distensibility, % | 1.76 ± 1.24 | 1.74 ± 1.3 | 0.65 |
| DAo max area, $cm^2$ | 474.68 ± 97.47 | 447.19 ± 83.32 | < 0.05 |
| DAo min area, $cm^2$ | 418.65 ± 92.13 | 396.29 ± 78.01 | 0.14 |
| DAo distensibility, % | 2.34 ± 1.43 | 2.17 ± 1.29 | 0.13 |
| Circumferential early PDSR, 1/s | 2.36 ± 0.71 | 2.02 ± 0.6 | < 0.001 |
| Radial early PDSR, 1/s | -5.94 ± 1.78 | -5.21 ± 1.54 | < 0.001 |
| Longitudinal early PDSR, 1/s | 1.68 ± 0.57 | 1.41 ± 0.43 | < 0.001 |

**Supplementary Table 3. Patient characteristics and CMR-derived measurements by genotype in patients with an EF $\geq$ 50%**. Categorical data - number of patients (relative frequency); Continuous data - mean ± standard deviation. BSA, body surface area; CMR, cardiac magnetic resonance imaging; DBP, diastolic blood pressure; EDV, end-diastolic volume; EF, ejection fraction; ESV, end-systolic volume; LA, left atrial; LV, left ventricular; LVM, left ventricular mass; RA, right atrial; RV, right ventricular; SBP, systolic blood pressure; WT, wall thickness; LVEDV, left ventricular end-diastolic volume; LVESV, left ventricular end-systolic volume; LVCO, left ventricular cardiac output; LVEF, left ventricular ejection fraction; RVEDV, right ventricular end-diastolic volume; RVESV, right ventricular end-systolic volume; RVEF, right ventricular ejection fraction; LAV, left atrial volume; LAEF, left atrial ejection fraction; RAV, right atrial volume; RAEF, right atrial ejection fraction; AAo, ascending aorta; DAo, descending aorta; PDSR, peak diastolic strain rate; GLS, global longitudinal strain; GRS, global radial strain; GCS, global circumferential strain.

**Supplementary Figures 1 - 5**

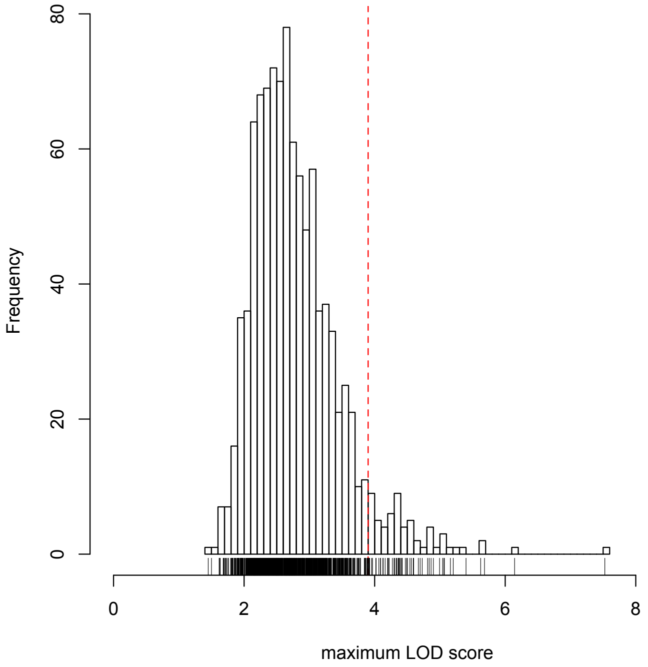


**Supplementary Figure 1. Permutation testing of quantitative trait loci in rats.** Distribution of LOD scores from 1000 permutation tests given the F2 intercross dataset showing that a LOD score of 3.9 will only be exceeded 5% by chance.

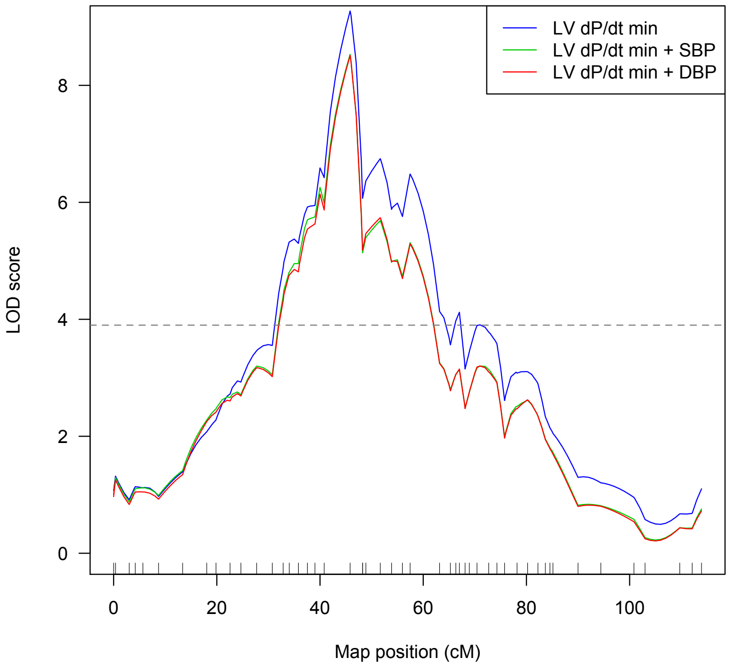


**Supplementary Figure 2. Effect of covariates on LOD scores.** QTL significance thresholds remain unaffected after taking into account the clinically important covariate effects of systolic and diastolic blood pressure on LV dP/dt$_{min}$.

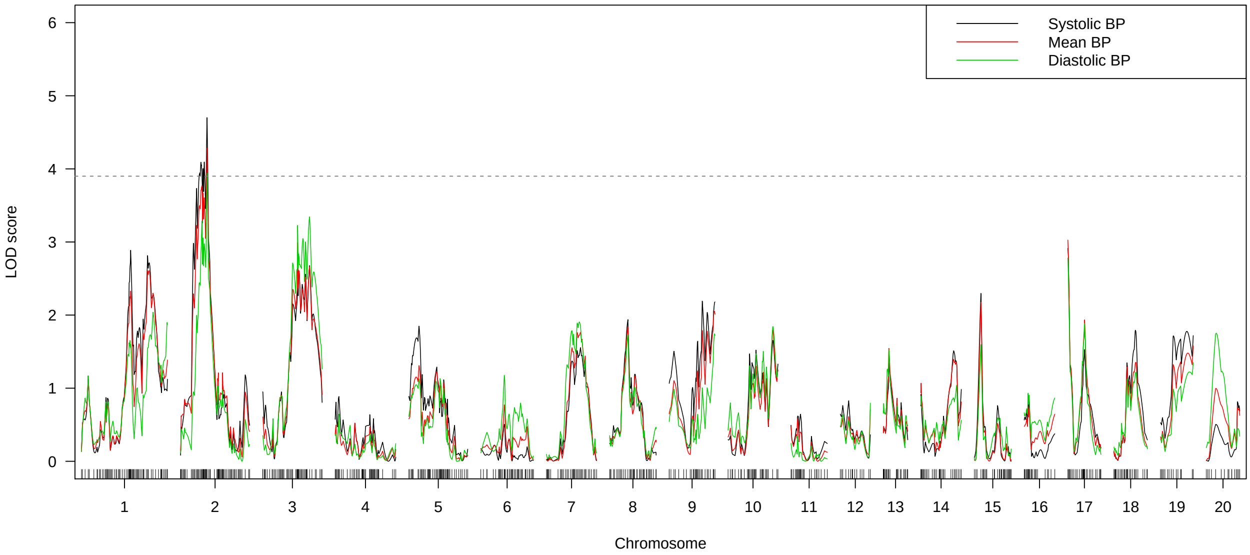


**Supplementary Figure 3. Effect of covariates on QTLs**. QTL significance thresholds remain unaffected after taking into account the clinically important covariate effect of SBP and DBP on LV dP/dtmin.

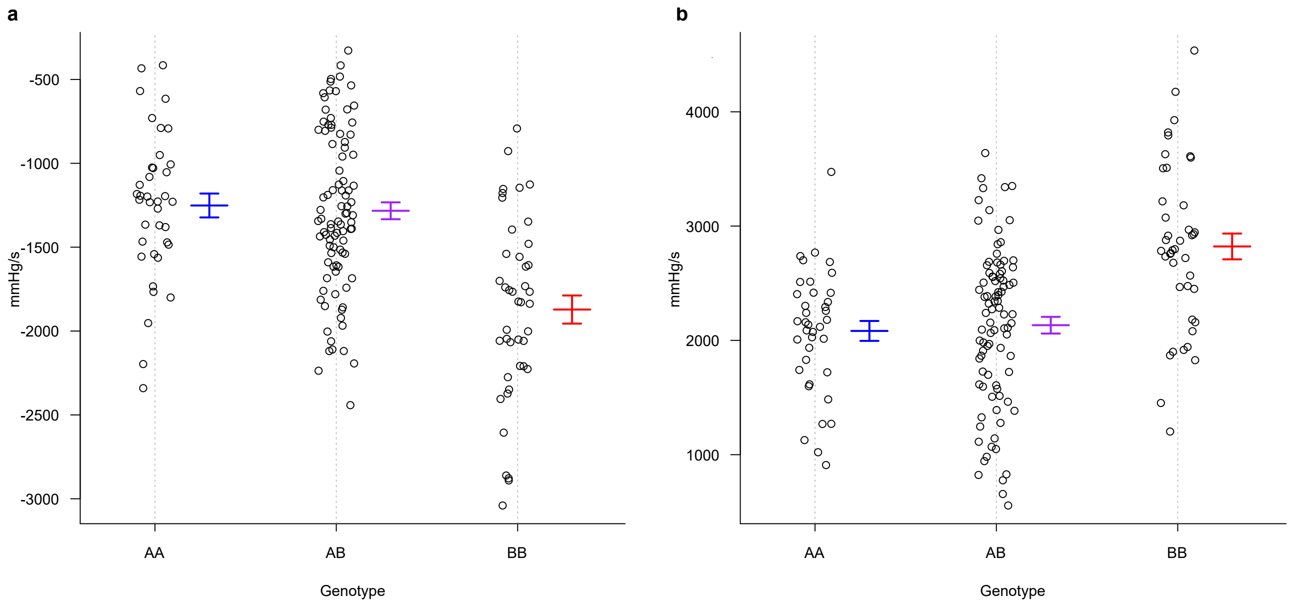


**Supplementary Figure 4. Titin alleles.** AA and BB refer to SHR and BN alleles at the locus respectively with AB referring to the heterozygous allele. BN allele (BB) is associated with better left ventricular **a.** relaxation ($dP/dt_{min}$) **b.** and contractility ($dP / dt_{max}$)

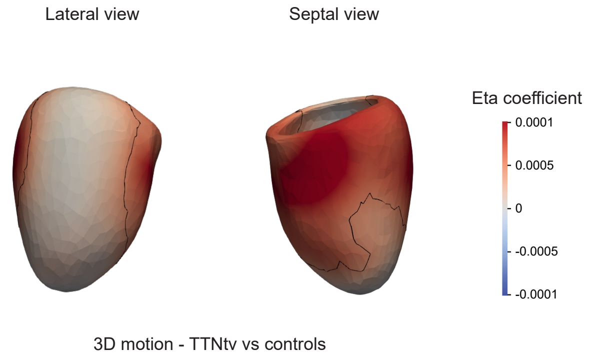


**Supplementary Figure 5. Mass multivariate analysis of cardiac motion phenotypes in three-dimensional left ventricle cardiac models.** To identify distributed patterns of heart activity associated with motion variations, mass multivariate linear regression was performed on vertex coordinates over time, showing a positive eta coefficient where TTNtv carrier status is associated with a greater motion coordinate variation. Sample size was 20,597 participants including 72 TTN+ individuals. Black outlines represent significant areas of differences between genotype status, with a significance level of $P \leq 0.01$.
